# Stroke prevalence among isolated systolic hypertension subjects in Indonesia

**DOI:** 10.1101/2021.11.27.21265906

**Authors:** Defi Pamelasari, Mahalul Azam, Arulita Ika Fibriana, Arief Rahadian, Muhamad Zakki Saefurrohim, Syed Mohamed Aljunid

## Abstract

**Introduction:** Hypertension has been known to be a decisive factor for stroke in the elderly; however, limited studies reported stroke risk factors in subjects with isolated systolic hypertension (ISH).

**Methods:** This cross-sectional survey was conducted using Riskesdas 2018 secondary data. Subjects aged 55 years who had a systolic blood pressure ≥140 mmHg and diastolic < 90 mmHg were included as the study sample. According to the National Institute of Health Research and Development, the Ministry of Health, Republic of Indonesia, all study variables were measured using household and individual questionnaires. The data were analyzed using the chi-square test and Fisher’s test.

**Results:** Of 3159 subjects with ISH 8.3% had a stroke, the risk factors that had a significant relationship with the prevalence of stroke (p<0.005) were gender (1,790; 1,420-2,256), smoking habits (1,645; 1,291-2,096), physical and mental stress (2,080; 1,618-2,673), area of residence (1,720; 1,331-2222), and education level (0.656; 0.515-0.835).

**Conclusion:** Prevalence of stroke among ISH in Indonesia was 8.3%. Female with smoking habits, had mental and physical stress, liviing in urban area, and had low level eduatcion were associated with the status of stroke in ISH subjects in Indonesia.

## INTRODUCTION

Stroke is the cause of death of the second and the cause of disability, the third in the whole world (1–3). During the past 15 years, the average stroke incidence has increased and reasons of death in low and medium state income (4). Indonesia occupied the order of the first in Asia with the number of cases of stroke most (4,5) and became the cause of death number one who killed 328.5 thousand people (21.2% of total deaths) in the year 2012 (6).

Studies on stroke risk factors have been widely reported, especially the incidence of stroke in subjects with diabetes mellitus (7) and hypertension (8). However, limited studies were reported on people with isolated systolic hypertension (ISH). Based on Framingham Heart Study data, the normotensive who reach the age of 65 years had a risk of 90% for the rest of life affected by hypertension (especially subtype systolic) if they lived 20 to 25 years longer (9). With the growing population aging too fast in the U.S., the prevalence of hypertension, especially ISH, is expected to increase substantially (10). Data are shown ISH occurs in 1% of the population aged 55 years in the AS, 5% at age 60 years, 12.5% at the age of 70 years, and 23.6% at age 75-80 years, especially on women (11).

ISH is defined as systolic blood pressure ≥140 mmHg and diastolic blood pressure <90 mmHg (12). ISH is a form of hypertension that most commonly occurs in the elderly (10,13). The phenomenon of an increase in systolic blood pressure in the elderly is believed to result from the secondary factors of risk that can be modified and changed in pathophysiological aging (10,11). The study reported that ISH is a risk factor of stroke prevalence (11). Limited studies reported the prevalence of stroke among patients with ISH. We aimed to explore the prevalence of stroke among ISH and the related factors in Indonesia.

## METHOD

The current study was a cross-sectional study. The sample study was a national health survey (riset kesehatan dasar=RISKESDAS) 2018. The sample in the study was the respondents who meet the criterion of ISH (systolic blood pressure ≥140 mmHg and diastolic blood pressure <90 mmHg), who were aged ≥55 years. Stroke determined based on the questionnaire delivered, with the previous diagnosed by a doctor. Other parameters and have the data complete the appropriate variables are examined, i.e., the information on the age, type of sex, the habit of smoking, consumption of alcohol, obesity, mental stress and physical status of the work, the area were living, level of education, consumption fruit, and vegetables. The data was acquired, processed, and analyzed using the program IBM SPSS Statistics 26. The test statistic used was the chi-square test and Fisher test because the type of data used was categorie, with a hypothetical comparative. The complete procedures of the research protocols can be found in the RISKESDAS data report (14) and our previous study(15).

## RESULTS

This study has acquired a total of 75,765 subjects with hypertension and completed data of the blood pressure. Of them, 3,159 subjects were categorized as ISH, aged ≥55 years old and included in the final analysis. The national prevalence of stroke in patients with systolic hypertension was 8.3%, which is shown in Table 1

**Table 1.** Characteristics of Study Subjects

| **Characteristics** | **N**<br>**(Total=3,159)** | **%** |
| --- | --- | --- |
| **Age** |  |  |
| 55-60 Years | 799 | 25.3 |
| 61 Years | 2360 | 74.7 |
| **Gender** |  |  |
| Men | 1061 | 33.6 |
| Women | 2098 | 66.4 |
| **Smoking** |  |  |
| Yes | 756 | 23.9 |
| No | 2403 | 76.1 |
| **Alcohol consumption** |  |  |
| Yes | 39 | 1.2 |
| No | 3120 | 98.8 |
| **Obesity** |  |  |
| Yes | 800 | 25.3 |
| No | 2359 | 74.7 |
| **Physical and mental stress** |  |  |
| Yes | 513 | 16.2 |
| No | 2646 | 83.8 |
| **Job status** |  |  |
| Unemployee | 1675 | 53 |
| Employee | 1484 | 47 |
| **Residential area** |  |  |
| Urban | 1837 | 58.2 |
| rural | 1322 | 41.8 |
| **Level of education** |  |  |
| Low (Under high school) | 2338 | 74 |
| High (High school or higher) | 821 | 26 |
| **Consumption of fruits and vegetables** |  |  |
| Not adequate | 2631 | 83.3 |
| Adequate | 528 | 16.7 |
| **Stroke** |  |  |
| Yes | 261 | 8.3% |
| No | 2,898 | 91.7% |

Table 1 also shows the characteristics of the respondents are aged ≥61 years (74.7%) and between 55-60 years (25.3%). Regarding gender, female (33.6%) was lower than male (66.4%). Most study respondents did not smoke (76.1%) and did not consume alcohol (98.8%). In addition, the characteristics of the sample were also dominated in non obese subjects (74.7%) and did not experience with mental and physical stress (83.8%). The frequency of respondents who did not work was 53%, live in urban areas was 58.2%, had low education was 74%, and less consume pf fruit and vegetables were 83.3%.

Tabel 2 shows the results of the bivariate analysis of factors associated with the occurrence of stroke in patients with ISH. that there was a relationship that significant between types of sex (p value = 0.000; RP = 1.790; 95% CI = 1.420 to 2.256), the habit of smoking (p value = 0,000, RP=1,645; 95% CI=1,291-2,096), physical and mental stress (p value = 0,000; RP=2,080; 95% CI=1,618-2,673), area of residence (p value = 0,000; RP=1,720; 95% CI=1,331-2.222), and education level (p value = 0,001; RP=0,656; 95% CI=0,515-0,835), with the incidence of stroke in ISH patients in Indonesia. While the variables that have no relation with the incidence of stroke was age (p value = 1.000; RP = 1.000; 95% CI = 0.765 to 1.306), the habit of drinking alcohol (p value = 0.768, RP = 0.618; 95% CI = 0.159 -2,395), obesity (p value = 0.952; RP = 1.018; 95% CI = 0.781-1.329), employment status (p value = 0.785; RP = 1.041; 95% CI = 0.824-1.314), and consumption of fruit and vegetables (p value = 1,000; RP=0.990; 95% CI=0.726-1.350).

**Table 2.** Subjects’ characteristics based on of Stroke status

| Characteristics | Stroke |  | P-value | Prevalence Ratio<br>(95% CI) |
| --- | --- | --- | --- | --- |
|  | Yes<br>n(%) | No<br>n(%) |  |  |
| **Age** |  |  |  |  |
| 55-60 years old | 66 (8.3) | 733 (91.7) | 1,000 | 1,000 (0.765-1.306) |
| 61 years old | 195 (8.3) | 2165 (91.7) |  |  |
| **Gender** |  |  |  |  |
| Man | 124 (11.7) | 937 (88.3) | 0.000 | 1,790 (1,420-2.256) |
| Woman | 137 (6.5) | 1961 (93.5) |  |  |
| **Smoking** |  |  |  |  |
| Yes | 89 (11.8) | 667 (88.2) | 0.000 | 1,645 (1,291-2,096) |
| No | 172 (7.2) | 2231 (92.8) |  |  |
| **Alcohol consumption** |  |  |  |  |
| Yes | 2 (5,1) | 37 (94.9) | 0.768 | 0.618 (0.159-2.395) |
| No | 259 (8.3) | 2861 (91.7) |  |  |
| **Obesity** |  |  |  |  |
| Yes | 67 (8.4) | 91.6 (91.6) | 0.952 | 1.018 (0.781-1.329) |
| No | 194 (8.2) | 91.8 (91.8) |  |  |
| **Physical and mental stress** |  |  |  |  |
| Yes | 75 (14.6) | 438 (85.4) | 0.000 | 2,080 (1,618-2,673) |
| No | 186 (7.0) | 2460 (93) |  |  |
| **Employment Status** |  |  |  |  |
| Employee | 141 (8.4) | 1534 (91.6) | 0.785 | 1.041 (0.824-1.314) |
| Unemployee | 120 (8,1) | 1364 (91.9) |  |  |
| **Residence** |  |  |  |  |
| Urban | 184 (10) | 1653 (90) | 0.000 | 1,720 (1,331-2,222) |
| Rural | 77 (5.8) | 1245 (94.2) |  |  |
| **Level of education** |  |  |  |  |
| Low (Under high school) | 170 (7.3) | 2168 (92.7) | 0.001 | 0.656 (0.515-0.835) |
| High (High school or higher) | 91 (11.1) | 730 (88.9) |  |  |
| **Consumption of fruits and vegetables** |  |  |  |  |
| Not adequate | 217 (8.2) | 2414 (91.8) | 1,000 | 0.990 (0.726-1.350) |
| Adequate | 44 (8.3) | 484 (91.7) |  |  |

## DISCUSSION

This study found the national prevalence of stroke in subjects with ISH was 8.3%. Age category did not significantly different among them. Previous study reported that age is one of the factors that influence the incidence of stroke (16). Patients with ISH found the respondents were dominated in female. Women, especially in elderly can lead an increase the incidence of stroke (17), the other studies alos reported that severity of stroke was also worse in women(18,19). The other complications of hypertension were also more prevalent in women than men (20).

Statistical tests show a significant relationship between smoking habits and the incidence of stroke in ISH patients in Indonesia. It was in line with several studies that say that smoking behavior has a significant relationship to the incidence of stroke (21,22). Cigarettes will affect the incidence of atherosclerosis, especially in the brain’s blood vessels, as a trigger for stroke (23)

Most of respondents had no habit in alcohol consumption, i.e. 98.8%. This study also found that there was no significant relationship between the consumption of alcohol and the incidence of stroke. Other study reported that alcohol consumption can increase the risk of stroke (24,25), limited subjetc that dominated by ISH in elderly in this study may influence the result (26).

Physical and mental stress had a relationship with the incidence of stroke among ISH subjects in Indonesia. Stress can stimulate the release of adrenaline and stimulate the heart to beat more rapidly and more robust, resulting in not stable blood pressure (27,28).

Residence type and level of education show a significant relationship with the incidence of stroke among ISH patients in Indonesia. Study in China stated that the burden of stroke in China has increased over the past 30 years and remains very high in the rural area (29). Other study stated that education leve, sex, lower level of education had a status of worse and higher occurance in severity and mortality (30). Low levels of education was significantly associated with an increased risk of death, stroke, recurrent, and the incidence of cardiovascular after stroke ischemic, apart from factors of risk are defined (31).

## CONCLUSION

Prevalence of stroke among ISH in Indonesia was 8.3%. Female with smoking habits, had mental and physical stress, liviing in urban area, and had low level eduatcion were associated with the status of stroke in ISH subjects in Indonesia.

## Data Availability

The data used to support the findings of this study are available from the corresponding author Mahalul Azam upon request through the email address.

## Acknowledgement

This study was supported by the Faculty of Sports Science, Universitas Negeri Semarang, Indonesia [Grant ID: 30.9.6/UN37/PPK.4.6/2021]. The RISKESDAS (Riset Kesehatan Dasar); a five-annual national basic health survey is conducted and supported by the National Institute of Health Research and Development (NIHRD), Ministry of Health, the Republic of Indonesia. The manuscript was prepared using a limited access data set obtained from the NIHRD and does not reflect the opinions or views of RISKESDAS and NIHRD. The authors thank the RISKESDAS investigators for granting permission to use their data set for the current study. The protocol and reports of the RISKESDAS is published on https://www.litbang.kemkes.go.id/laporan-risetkesehatan-dasar-riskesdas//

## Ethical Clearance

Ethical clearance for the RISKESDAS 2018 study was obtained from the Ethics Committee, the National Institute of Health Research and Development (NIHRD), the Ministry of Health, Republic of Indonesia

All necessary patient/participant consent has been obtained and the appropriate institutional forms have been archived.

